# Multicluster measles outbreak with a substantial proportion of modified cases in Tokyo, Japan, January–May 2026

**DOI:** 10.64898/2026.06.17.26355652

**Authors:** Yukari Nishikawa, Junko Kurita, Tamie Sugawara, Anna Matsuda, Ryuto Morikawa, Yuki Imokawa, Megumi Iwamura, Kaoru Suzuki, Shota Kodaira, Shouta Kikuchi, Aki Takahashi, Itaru Nishizuka, Mitsuo Kaku

## Abstract

Tokyo experienced a measles outbreak (260 cases) by late May 2026 despite elimination status. Adults aged 20–39 years were most affected, and 38% of cases were modified measles, increasing with prior vaccination. Although incidence rose until April, the effective reproduction number fell below 1, consistent with outbreak control. Multiple clusters were identified, but many cases lacked epidemiological links, suggesting modified measles is under-recognized. Intensive contact tracing and surveillance contributed to limiting transmission.

## Early 2026 measles situation in Tokyo and Europe

Japan was verified as having achieved measles elimination in 2015, and most subsequent cases were sporadic, import□related or in small clusters [1-3]. By late May 2026, however, Tokyo experienced a rapidly expanding urban outbreak despite elimination status [4]. Recent measles activity in Europe shows that the region remains at high risk for measles circulation despite long□standing two□dose programmes [5, 6]. This rapid communication describes the early epidemiology of the outbreak, the frequency and clinical profile of modified measles, and assesses outbreak control using the time□varying effective reproduction number (*R*(*t*)).

### Case definitions, ascertainment and *R*(*t*) estimation

We analysed all measles cases notified to the Tokyo Metropolitan Government between 1 January and 31 May 2026 under the Infectious Disease Control Law of Japan. Physicians report suspected measles cases to local public health centres, and these notifications are consolidated in the National Epidemiological Surveillance of Infectious Diseases System, which the Tokyo Metropolitan Government uses to monitor measles trends within the city.

Measles cases were classified into three categories. Those with the classic triad (rash, fever and catarrhal symptoms) and laboratory confirmation (measles virus RNA or measles-specific IgM detected), laboratory-confirmed classic measles; those with the triad but without laboratory confirmation, clinically diagnosed classic measles; and those not meeting the triad but with laboratory confirmation, laboratory-confirmed modified measles.

Key variables included demographics, vaccination history, exposure history, and laboratory/genotype results. Vaccination history was ascertained from maternal and child health handbooks and medical records. Cases were considered epidemiologically linked when a plausible source of infection was identified through contact tracing conducted by public health centres, and sporadic when no epidemiological link could be established despite investigation. A cluster was defined as two or more epidemiologically linked cases sharing a plausible transmission setting, such as a workplace or household, including onward transmission up to third□generation cases from the same setting, whereas epidemiological links also included infections acquired through contact with known cases in different settings.

The time-varying effective reproduction number, *R*(*t*), was estimated from the epidemic curve by symptom onset using a previously described method [7]. Missing onset dates were imputed from the onset-to-diagnosis interval distribution, and uncertainty was assessed by bootstrap resampling. Among cases with documented vaccination history, we compared the proportion of modified measles across vaccination dose categories using Fisher’s exact test and the Cochran–Armitage trend test, and we estimated relative risks with 95% confidence intervals for hospitalisation or complications. This study used surveillance data collected for public health purposes under Japanese law; institutional review board approval was not required. We followed the STROBE guideline[8].

Between 1 January and 31 May 2026, 260 measles cases were reported in Tokyo, with a median age of 25 years (IQR: 16.0–32.0); 171 cases were male (65.8%). The most affected age groups were 20–29 years (98/260; 37.7%), 30–39 years (55/260; 21.2%), and 10–19 years (52/260; 20.0%). Among all cases, 161 (61.9%) were classified as classic measles, 99 (38.1%) as modified measles (Table 1).

**Table 1.** Baseline characteristics of 260 measles cases, Tokyo, January–May 2026.

| Characteristic | n | % |
| --- | --- | --- |
| Sex |  |  |
| Male | 171 | 65. |
|  |  | 8 |
| Age (years) |  |  |
| 0-9 | 22 | 8.5 |
| 10-19 | 52 | 20.<br>0 |
| 20-29 | 98 | 37.<br>7 |
| 30-39 | 55 | 21.<br>2 |
| ≥40 | 33 | 12.<br>7 |
| Case type |  |  |
| Classic measles with laboratory confirmed | 159 | 61.<br>2 |
| Classic measles clinically diagnosed | 2 | 0.8 |
| Modified measles | 99 | 38.<br>1 |
| Genotype |  |  |
| D8 | 221 | 85.<br>0 |
| B3 | 26 | 10.<br>0 |
| Undetermined | 13 | 5.0 |
| Epidemiological link |  |  |
| Probable | 119 | 45.<br>8 |
| No known link | 141 | 54.<br>2 |
| Cases in identified clusters | 97 | 37.<br>3 |
| Hospitalized | 17 | 6.5 |
| Complications | 6 | 2.3 |
| Death | 0 | 0 |
| International travel during incubation period | 21 | 8.1 |

The epidemic curve showed progressive growth from sporadic cases in January to a weekly peak of 55 cases in epidemiological week 17. From mid-April, *R*(*t*) remained below 1 despite high weekly case counts, suggesting progressively curtailed transmission with ongoing case detection (Figure 1).

**Figure 1.**
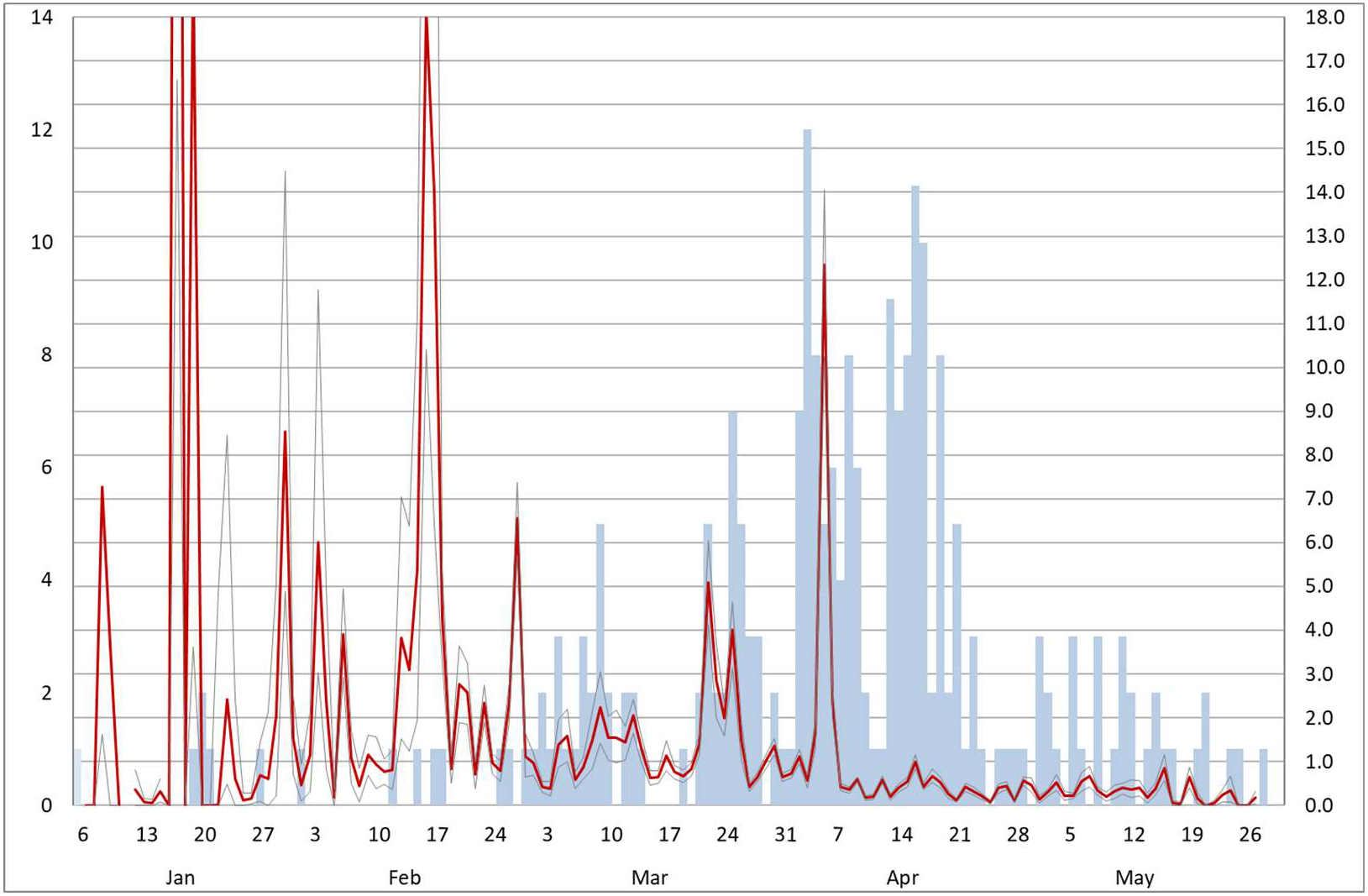
Estimated time-varying effective reproduction number and symptom onset up to 31 May 2026: dark blue bars indicate the number of reported cases by date of symptom onset (left axis), the red line shows the estimated effective reproduction number; *R*(*t*), and thin black lines represent the 95% confidence interval for *R*(*t*).

### Clinical presentation and role of vaccination

Vaccination history was documented for 132 of 260 cases (50.8%). Among cases with known vaccination history, the proportion of modified measles increased with the number of documented vaccine doses, from 4/22 (18.2%) in unvaccinated individuals to 13/28 (46.4%) in one-dose recipients and 47/82 (57.3%) in recipients of two or more doses. Overall, 47 of 99 modified measles cases occurred in people with at least two documented doses (Figure 2; Supplementary Table S2).

**Figure 2.**
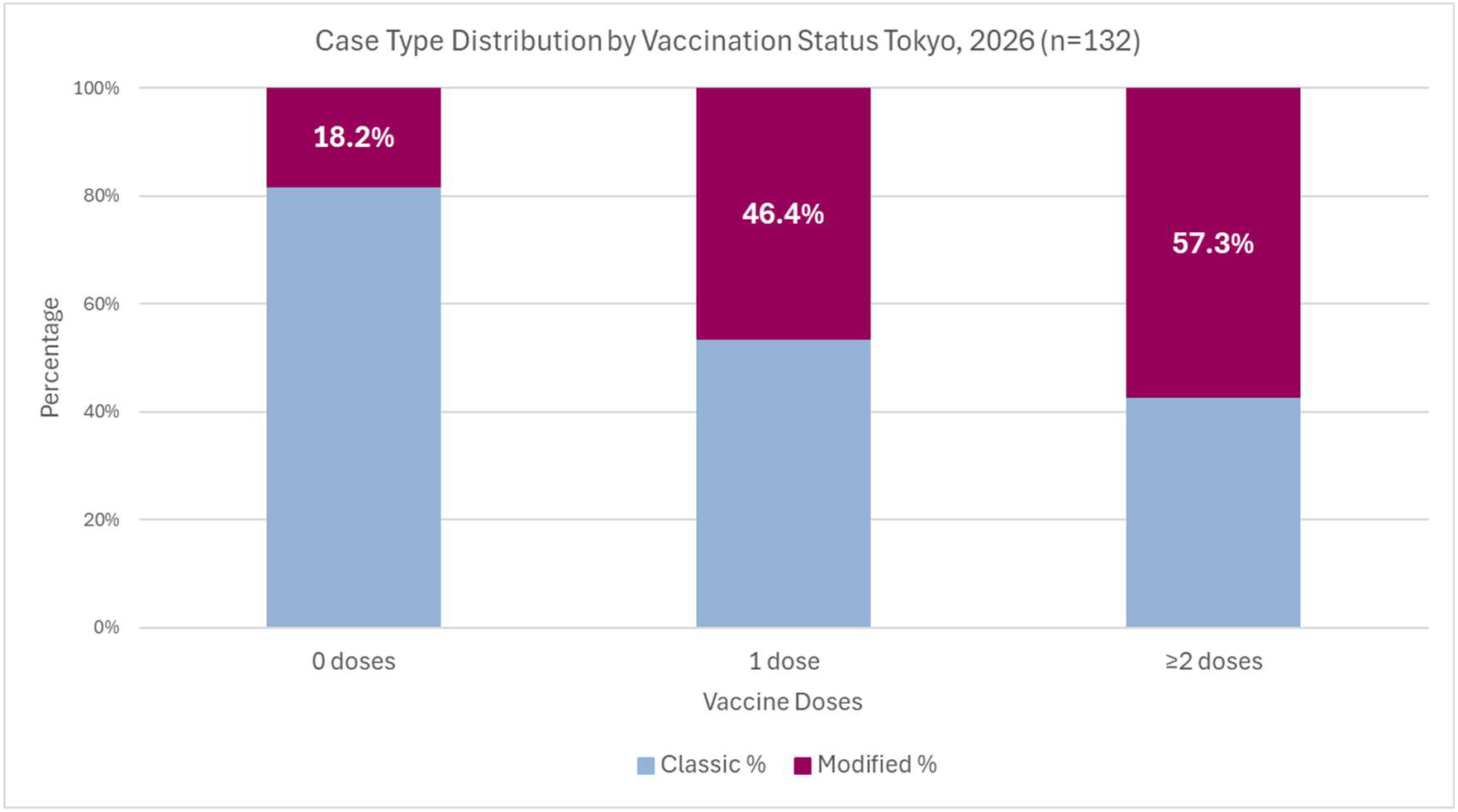
Proportion of modified measles among measles cases by number of documented measles-containing vaccine doses, Tokyo, 2026 (n=132): bars show the proportion of modified measles among unvaccinated, one□dose, and over two□dose recipients with known vaccination history.

Among cases with known vaccination history, hospitalisation or complications occurred in 5 of 22 unvaccinated cases (22.7%), 0 of 28 one-dose recipients, and 3 of 82 two-dose recipients (3.7%), suggesting a lower risk of severe outcomes among vaccinees (Supplementary Table S3).

## Discussion

A key finding was the high proportion of modified measles, particularly among vaccinees with documented prior doses. Among cases with known vaccination history, the proportion of modified measles increased with the number of documented doses, compatible with waning vaccine□derived protection in the absence of natural boosting [10]. Despite this shift towards modified disease, documented two-dose recipients were much less likely to require hospitalisation or experience complications than unvaccinated cases, suggesting substantial retained protection against severe clinical outcomes.

Ten epidemiological clusters comprising 97 of 260 cases (37.3%) were identified through integration of active epidemiological investigation data from individual public health centres. Overall, 53 of 97 cluster□associated cases (54.6%) were classified as modified measles, and 55 had documentation of two or more prior doses (56.7%). The two largest clusters occurred in school settings and involved 10 and 54 cases, respectively (Supplementary Table S4).

A second important finding was the divergence between the increasing proportion of cases without identified epidemiological links and declining *R*(*t*). One plausible explanation is that modified measles, because of its attenuated and non-specific presentation, is more likely than classic measles to be under-recognised and remain unlinked epidemiologically [7]. The identification of distinct D8- and B3-associated clusters through routine genotyping at the Tokyo Metropolitan Institute of Public Health and integration of epidemiological investigation data from public health centres identified additional transmission chains not apparent from routine notification data and supported timely interruption of transmission.

Together with the substantial proportion of cases without identified epidemiological links, this experience indicates that routine case-based surveillance alone is unlikely to capture all transmission chains in an elimination setting, and that intensive field investigation and systematic contact tracing may have contributed to constraining further spread, as reflected in the sustained fall of *R*(*t*) below 1 [11].

This outbreak shows that measles transmission can still occur in an elimination setting, but also that intensive surveillance, including molecular epidemiological investigations, and active contact tracing can reduce transmission rapidly. Consistent with this, *R*(*t*) fell below 1 by mid□April and remained there despite sustained case reporting.

The proportion of modified measles in this outbreak was higher than that reported in Yamagata in 2017 [12] and broadly consistent with concerns raised during the 2018 Okinawa outbreak and recent European outbreaks about breakthrough and modified presentations in highly vaccinated populations [7, 13]. These findings support heightened clinical suspicion for modified measles in vaccinated adolescents and adults, particularly in schools and other congregate settings, including in highly vaccinated urban areas in Europe.

This analysis has several limitations. Vaccination history was unknown for many cases. Vaccination was considered present only when confirmed in maternal and child health handbooks or other official records; thus, coverage was likely underestimated, particularly among adults in their 20s and 30s, in whom these handbooks were often unavailable or had been lost. Modified measles was defined operationally as cases not fulfilling the clinical triad of fever, rash, and catarrhal symptoms, which may not fully capture the clinical spectrum. The outbreak was ongoing at the time of analysis, and the findings reflect the situation as of late May 2026; final case counts and transmission patterns may change. Investigations into waning immunity breakthrough infections, and detailed cluster dynamics are ongoing, and a full surveillance report will be submitted.

## Data Availability

Aggregated data from the National Epidemiological Surveillance of Infectious Diseases (NESID) system, operated under the Infectious Diseases Control Law in Japan for data collection (Articles 12 and 14), expert analysis, and public reporting (Article 16), were used in this study. All data used in the analyses are publicly available as aggregated surveillance data from the NESID system and do not contain personally identifiable information.

